# Effects of return-to-office, public schools reopening, and vaccination mandates on COVID-19 cases among municipal employee residents of New York City

**DOI:** 10.1101/2022.10.17.22280652

**Authors:** Sharon K. Greene, Bahman P. Tabaei, Gretchen M. Culp, Alison Levin-Rector, Nishant Kishore, Jennifer Baumgartner

## Abstract

**Objective:** On September 13, 2021, teleworking ended for New York City municipal employees, and Department of Education (DOE) employees returned to reopened schools. On October 29, COVID-19 vaccination was mandated. We assessed these mandates’ short-term effects on disease transmission.

**Methods:** Using difference-in-difference analyses, we calculated COVID-19 incidence rate ratios (IRR) among residents 18–64 years-old by employment status pre- and post-policy implementation.

**Results:** IRRs post-(September 23–October 28) vs. pre-(July 5–September 12) return-to-office were similar between office-based City employees and non-City employees. Among DOE employees, the IRR after schools reopened was elevated 28.4% (95% CI: 17.3%–40.3%). Among City employees, the IRR post-(October 29–November 30) vs. pre- (September 23– October 28) vaccination mandate was lowered 20.1% (95% CI: 13.7%–26.0%).

**Conclusions:** Workforce mandates influenced disease transmission, among other societal effects.

---

To encourage social distancing while ensuring continuity of operations during the COVID-19 outbreak, the New York City (NYC) Department of Citywide Administrative Services (DCAS) issued a temporary telework policy on March 13, 2020 for eligible municipal employees of the City of New York (hereafter referred to as City employees).^1^ Following COVID-19 vaccination availability, the mayor directed all City employees to return to the office at least part-time by May 3, 2021^2^ and full-time by September 13, 2021,^3^ with protective measures in place including mandatory face coverings^4^ and proof of full vaccination or weekly negative PCR diagnostic tests.^5^ As justification for withdrawing the telework policy, a mayor’s spokesperson stated, “We know how to make workplaces safe, and public servants can deliver more for New Yorkers when they’re working together.”^3^

During the 2020–2021 academic year, a remote learning option was available to families with children in public schools.^6^ When the 2021–2022 academic year commenced on September 13, 2021, public schools returned to full-time, in-person instruction.^7^ Staff and students were required to wear face coverings while on school property.^8^ As with all City employees, unvaccinated teachers and staff were required to test weekly.^9^

By order of the Commissioner of Health and Mental Hygiene, COVID-19 vaccination was required for Department of Education staff effective October 1, 2021^10^ and more generally for all City employees effective October 29, 2021.^11^ This order cited a federal executive order stating, “It is essential that Federal employees take all available steps to protect themselves and avoid spreading COVID-19 to their co-workers and members of the public. The [U.S. Centers for Disease Control and Prevention] CDC has found that the best way to do so is to be vaccinated.”^12^

COVID-19 outbreaks have been documented across various worksite settings.^13-15^ In California, where employer reporting of workplace COVID-19 outbreaks was mandated, the public administration sector (including correctional, police, and fire services) had the highest incidence of reported workplace outbreaks through August 2021.^16^ Given the importance of evaluating the effectiveness of public health interventions for COVID-19,^17-19^ we assessed whether the mandates achieved their objectives with respect to minimizing workplace transmission among NYC employees. NYC has the largest municipal workforce in the U.S.,^20^ and the City of New York is the largest single employer in the New York metropolitan area.^21^

Specifically, we used a quasi-experimental study design to compare COVID-19 case rates among City employees relative to other NYC residents of working age. We assessed whether the September 13, 2021 return-to-office mandate and reopening of public schools was not associated with a relative increase in cases and whether the October 29, 2021 vaccination mandate was associated with a relative decrease in cases.

## METHODS

### Study Population

The study population was NYC residents of working age, 18–64 years-old. We defined City employees as persons appearing on lists of municipal employees as of both July 8, 2021 and November 29, 2021, provided by DCAS under a data use agreement with the NYC Department of Health and Mental Hygiene (DOHMH). The DCAS lists were stored on a secure server with access limited to authorized DOHMH study personnel. For the denominator for City employee case rates (N = 212,953), we restricted to persons who, as of both dates, worked at the same City agency, were 18–64 years-old, and did not live outside of NYC. As NYC residency status could not be determined for 33,854 uniformed Police Department employees with a worksite instead of residential address on the DCAS lists, we assumed 48% were NYC residents.^22^ Non-City employees were defined as all other NYC residents 18–64 years-old (N = 5,001,460), approximated as the 2020 intercensal population estimate^23^ minus the number of 18–64 year-old NYC residents appearing on either DCAS list except the 52% of uniformed Police Department employees assumed to be non-NYC residents. Persons appearing on only one of the two DCAS lists were excluded, as the purpose was to assess effects of mandates on COVID-19 case trends in closed cohorts.

The primary outcome of interest was diagnosis with a confirmed or probable case of COVID-19, per the national surveillance case definition.^24^ Cases were ascertained primarily through electronic laboratory reporting through the New York State Electronic Clinical Laboratory Reporting System.^25^ Patients whose address at time of report indicated residence in a nursing home, adult care facility, jail, or prison were excluded. Symptom status was ascertained by routine interview, e.g., for contact tracing, and we classified patients as symptomatic if they met clinical criteria for COVID-19–like illness, per the surveillance case definition.^24^ Case data were extracted from the DOHMH COVID-19 surveillance database (Maven® Disease Surveillance and Outbreak Management System; Conduent, Florham Park, New Jersey) on February 15, 2022.

We primarily assessed trends in cases, as opposed to hospitalizations or deaths, because the stated rationale for the mandates, as above, emphasized making workplaces safe and avoiding spreading COVID-19 to co-workers; additionally, the study population excluded ≥65 year-olds, the group at highest risk for severe illness. However, in a secondary analysis to assess the first three months after the municipal employee vaccination mandate was implemented, we considered COVID-19 hospitalizations as an additional outcome of interest. These were ascertained by importing and matching data from supplemental systems, as previously described^26^ and defined as patients having a positive SARS-CoV-2 test within 14 days before or 3 days after hospital admission. Because COVID-19 hospitalization risk increases with age,^27^ we compared the age distribution of City and non-City employees as of November 27, 2021, and assessed hospitalization trends in the overall 18–64 year-old study population as well as restricting to 50–64 year-olds.

### Matching Employee and Case Data

Employee lists and cases were geocoded with version 22A of the NYC Department of City Planning’s Geosupport geocoding software,^28^ implemented in R through C++ using the Rcpp package.^29^ Addresses that failed to geocode were then cleaned using a string searching algorithm performed against the Department of City Planning’s Street Name Dictionary and Property Address Directory.^28^ Addresses that still failed to geocode after cleaning were then run through a United States Postal Service verification service^30^ for further cleaning and to flag addresses outside of NYC.

We used three sets of geocoder outputs. First, the building classification code^31^ was used to flag non-residential addresses. Employees of certain City agencies (Police Department, Department of Corrections, and Department of Investigations) commonly self-reported to DCAS their work instead of home address due to security concerns. We matched cases diagnosed among such employees with laboratory reports of SARS-CoV-2 molecular and antigen tests, regardless of test date and result. This match used fuzzy matching on first and last name, exact matching on birth date, and the ordering facility name as available to confirm City employee status because Police Department employees frequently accessed tested through their employer. Cases diagnosed among employees with a valid NYC residential address on any laboratory report were considered NYC residents and study-eligible; otherwise, cases were excluded from analysis because employee residency in NYC could not be assumed.

Secondly, geocoder outputs of the tax lot identification number (borough-block-lot) and building identification number were used in record matching, as described below. We prioritized these standardized geocoder outputs for matching over address fields to minimize problems with address data entry and missingness. Thirdly, the United States Census Bureau block was converted using Topologically Integrated Geographic Encoding and Referencing data^32^ to 2010 ZIP Code Tabulation Areas, which were in turn used to determine United Hospital Fund (UHF) neighborhood (N = 42), a geography that aggregates adjoining ZIP code areas of similar characteristics to approximate community districts.^33,34^ UHF neighborhoods were used to assess geographic representativeness of NYC employees compared with the general population, as described below.

To assign City employment status to each COVID-19 case, we used a multistep, deterministic, one-to-many hierarchical record matching algorithm. We linked records using a series of “keys” consisting of exact character correspondence between variables.^35^ We constructed 17 matching keys in the DCAS and surveillance case linelists using combinations of first name, middle name, last name, date of birth, borough-block-lot, and building identification number; see text, Supplemental Digital Content 1, which shows the steps for dataset cleaning, standardization, and reformatting, as well as the matching keys.

### Geographic Representativeness of NYC Employees

To assess support for the assumption that City employees and non-City employees residing in NYC were exposed to the same underlying epidemic trends, we compared the geographic distribution of City employees with that of the general population 18–64 years-old by UHF neighborhood of residence. This analysis included employees who had the same residential UHF neighborhood in DCAS lists from both July 8 and November 29, 2021, as well as employees whose residential address was not available in either DCAS list but was available through matching to case or laboratory testing data.

### Difference-in-Difference Analysis

Difference-in-difference analyses are used to compare group means of an outcome before and after policy implementation. The counterfactual is based on a similar comparator group that was not subject to the policy intervention.^36^ Described as a generalized linear model, the statistical components are:

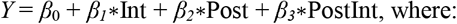

*Y* is the outcome of interest, i.e., COVID-19 cases per 100,000 person-days, accounting for pre- and post-implementation periods of unequal durations. Int is a dummy variable that is 1 if the observation occurred in the group that experienced the intervention (City employees) or 0 otherwise (non-City employees). Post is a dummy variable that is 1 if the observation occurred temporally after the policy implementation and 0 otherwise. PostInt is a product of Int and Post and is a dummy variable that is 1 if the observation occurred temporally after the policy implementation in the intervention group or 0 otherwise.

The four coefficients can be interpreted as: *β*_0_ is the mean outcome in the non-intervention group (non-City employees) in the pre-intervention period (pre-mandate). *β*_*1*_ is the difference in mean outcome in the intervention group (City employees) in the pre-intervention period compared with the mean outcome in the non-intervention group in the pre-intervention period. *β*_*2*_ is the difference in the mean outcome in the non-intervention group in the post-intervention period (post-mandate) compared with the mean outcome in the non-intervention group in the pre-intervention period. *β*_*3*_ it the measure of interest, or the difference-in-difference, i.e., the additional difference in the mean outcome in the intervention group in the post-intervention period compared with the mean outcome in the intervention group in the pre-intervention period when compared with the difference defined for *β*_*2*_.

We used a Poisson link in the generalized linear model, with a log-transformed offset term for person-time at risk. Therefore, the coefficients of interest are interpreted as differences in the log incidence rates in the contrasts of interest described above. We exponentiated these estimates and their respective confidence intervals (CI) to calculate the incidence rate ratios of the contrasts described above. For example, exponentiating *β*_*2*_ yielded the ratio of the COVID-19 incidence rate in non-City employees post-mandate compared with pre-mandate. To estimate the incidence rate in City employees post-mandate compared with pre-mandate, we summed the *β*_*2*_ and *β*_*3*_ estimates before exponentiating them and extracted the variance-covariance matrices of these covariates to calculate the standard error and subsequent 95% CI.

### Return-to-Office Mandate and Reopening of Public Schools (September 13, 2021)

To estimate the effect of the return-to-office mandate on COVID-19 transmission, we compared changes in COVID-19 case rates from pre-to post-mandate implementation periods for office-based City employees relative to other working-age adults. Office-based City employees (N = 80,454) were defined by excluding City employees who were unlikely to have worked remotely during the pre-implementation period, i.e., uniformed employees of the Department of Corrections, Department of Sanitation, Fire Department, and Police Department; Department of Education employees; and employees with a civil service title description of city seasonal aide. To estimate the effect of reopening public schools, we restricted to Department of Education employees (N = 97,879).

We defined the pre-implementation period as diagnoses during July 5–September 12, 2021, i.e., the 10 weeks prior to the mandate and coinciding with the increasing slope of NYC’s third epidemic wave.^37^ If the return-to-office mandate or reopening of public schools were associated with increased disease transmission, then it would take time for employees to become infected and develop symptoms. Assuming a median incubation period of 5 days,^38^ we imposed a washout period (September 13–22, 2021) of two incubation periods for worksite transmission to occur. Laboratory-based testing availability in NYC during this period was widespread, so we did not build in extra time from symptom onset to testing. Before students returned on September 13, 2021, Department of Education staff began reporting to schools on August 30 or September 9, 2021, depending on their job title.^39^ The post-implementation period was defined as September 23–October 28, 2021, ending prior to the vaccination mandate and coinciding with a citywide decline in cases.^37^ The Delta variant predominated throughout these pre- and post-implementation periods.^40^

We considered the interventions to be effective with respect to disease transmission if the change in COVID-19 case rates using a non-inferiority test^41^ from the pre-to post-implementation periods was not larger for office-based City employees or Department of Education employees than the comparison group by a pre-specified margin. This margin is a policy decision as to what COVID-19 case rate is considered acceptable in the workplace as a trade-off for the benefits of in-person work. The policies did not define a “safe” level of transmission. At the time, CDC defined a low community transmission level as <10 new cases per 100,000 persons in the past 7 days.^42^ Thus, we *a priori* defined an acceptable margin consistent with “low” worksite transmission as <10 excess average weekly cases per 100,000 employees during the post-implementation period.

### Vaccination Mandate (October 29, 2021)

The vaccination mandate was layered on top of the return-to-office mandate. We defined the pre-implementation period for the vaccination mandate to be the same as the post-implementation period for the return-to-office mandate (September 23–October 28, 2021). On October 29, 2021, the weekly testing option was rescinded, and City employees were required to have received at least one COVID-19 vaccine dose.^11,43^ The post-implementation period for the vaccination mandate was defined as October 29–November 30, 2021, ending prior to identification of the first confirmed U.S. case of infection with the Omicron SARS-CoV-2 variant on December 1, 2021.^44^ Cases increased rapidly in NYC throughout December,^37^ with Omicron constituting 75% of sequencing results by week ending December 18, 2021.^40^ Given reduced vaccine effectiveness against infection with the Omicron variant,^45^ we did not necessarily expect the vaccination mandate (which included no requirement for a booster dose) to strongly reduce disease transmission after Omicron emergence. In addition, COVID-19 vaccination was required for the general workforce in NYC effective December 27, 2021,^46^ after which effects of the vaccination mandate on COVID-19 cases among City employees could be biased toward the null. Nevertheless, in a secondary analysis, we defined an extended post-implementation period as October 29, 2021–January 31, 2022 to assess COVID-19 cases and hospitalizations diagnosed during the first three months after the municipal employee vaccination mandate was implemented, including during the large Omicron (BA.1) epidemic wave.

We compared changes in COVID-19 case rates over time between City employees and other working-age adults. We considered the vaccination mandate to be effective with respect to disease transmission if the change in COVID-19 case rates from the pre-to post-implementation periods was statistically significantly lower in City employees than in the comparison group, controlling for differences between groups in the pre-implementation period.

Compliance with the vaccination mandate at the time of implementation varied across agencies. The reported percentage of an agency’s workforce (including City employees who were not NYC residents) with at least one COVID-19 vaccine dose as of October 30, 2021 ranged from 60% at the Department of Corrections to 100% at the Landmarks Preservation Commission.^47^ We grouped employees of agencies with higher vs. lower vaccination coverage, defined as ≥96% vs. <90% with at least one vaccine dose as of October 30, 2021.^47^ If the vaccination mandate had been effective in reducing disease transmission, then in a difference-in-difference-in-difference (triple difference) analysis,^36^ employees of agencies with higher vaccination coverage might be expected to have experienced greater reductions in COVID-19 case rates from the pre-to post-implementation periods relative to other working-age adults than employees of agencies with lower vaccination coverage.

## RESULTS

Of 921,057 confirmed and probable COVID-19 cases diagnosed among community-dwelling NYC residents 18–64 years-old during July 5, 2021–January 31, 2022, 45,291 (4.9%) were among eligible City employees, and 863,459 (93.7%) were among non-City employees. The remaining 12,307 (1.3%) were excluded as ineligible (diagnosed in a person present on only one of the two DCAS lists; present on both DCAS lists but employed at different agencies; having a non-NYC address on one of the DCAS lists; or Police Department, Department of Corrections, or Department of Investigations employees using their work address and for whom NYC residency could not be assumed).

Within municipal employment categories, the percentage of case-patients known to have had COVID-19–like illness was similar across diagnosis periods (Table 1), suggesting that any changes in testing rates over time did not lead to large differences in the proportion of infections ascertained as cases. The percentage of case-patients known to have COVID-19–like illness was similar between City employees (87.1%) and non-City employees (84.9%).

**Table 1.** Percentage of COVID-19 cases among New York City residents 18–64 years-old with COVID-19–like illness, by diagnosis period and municipal employment status.

| Mandate | Period | Non-City employees<br>(N = 5,001,460) |  |  | City employees |  |  |  |  |  |  |  |  |
| --- | --- | --- | --- | --- | --- | --- | --- | --- | --- | --- | --- | --- | --- |
|  |  |  |  |  | All<br>(N = 212,953) |  |  | Office-based*<br>(N = 80,454) |  |  | Department of Education<br>(N = 97,879) |  |  |
|  |  | Cases | Cases with known symptom status (%) | Cases with known symptom status and COVID-19–like illness (%) | Case count | Cases with known symptom status (%) | Cases with known symptom status and COVID-19–like illness (%) | Cases | Cases with known symptom status (%) | Cases with known symptom status and COVID-19–like illness (%) | Case count | Cases with known symptom status (%) | Cases with known symptom status and COVID-19–like illness (%) |
| Pre-return to-office/<br>schools reopening | July 5–<br>September 12, 2021 | 73,113 | 56,771 (77.6%) | 48,532 (85.5%) | 3,677 | 3,040 (82.7%) | 2,646 (87.0%) | 1,128 | 969 (85.9%) | 844 (87.1%) | 1590 | 1,348 (84.8%) | 1,173 (87.0%) |
| Post-return to-office/schools reopening and pre-vaccination | September 23–<br>October 28, 2021 | 25,540 | 19,188 (75.1%) | 16,017 (83.5%) | 1,464 | 1,211 (82.7%) | 1,043 (86.1%) | 412 | 332 (80.6%) | 262 (78.9%) | 713 | 639 (89.6%) | 595 (93.1%) |
| Post-vaccination | October 29–<br>November 30, 2021 | 28,019 | 21,854 (78.0%) | 18,464 (84.5%) | 1,284 | 1,082 (84.3%) | 956 (88.4%) | 362 | 296 (81.8%) | 250 (84.5%) | 646 | 576 (89.2%) | 535 (92.9%) |
| Total |  | 126,672 | 97,813 (77.2%) | 83,013 (84.9%) | 6,425 | 5,333 (83.0%) | 4,645 (87.1%) | 1,902 | 1,597 (84.0%) | 1,356 (84.9%) | 2,949 | 2,563 (86.9%) | 2,303 (89.9%) |

\*Office-based City employees excluded uniformed employees of the Department of Corrections, Department of Sanitation, Fire Department, and Police Department; Department of Education employees; and employees with a civil service title description of city seasonal aide.

Of NYC residents of working age, 33.5% of City employees were 50–64 years-old, a slightly higher proportion than the 28.3% of non-City employees (see Figure, Supplemental Digital Content 3, which illustrates that that among NYC residents 18–64 years-old, City employees generally skewed older than non-City employees).

### Geographic Representativeness of NYC Employees

The residential geographic distribution of City employees was moderately well correlated with that of the general population (see Figure, Supplemental Digital Content 2, which illustrates that UHF neighborhoods where larger percentages of NYC employees resided generally also had larger percentages of the general population). The Pearson correlation coefficient across the 42 neighborhoods was 0.58 (*P*<0.0001). NYC employees were over-represented in the South Beach and Tottenville neighborhood of Staten Island (UHF neighborhood 504,^34^ with 6.7% of City employees and 2.2% of general population) and under-represented in West Queens (UHF neighborhood 402, with 3.5% of City employees and 5.5% of general population).

### Return-to-Office Mandate and Reopening of Public Schools

The Delta epidemic wave was waning in NYC when the return-to-office mandate was implemented and public schools reopened on September 13, 2021^37^ (Figure 1). Accordingly, the case rate per 100,000 person-days decreased during the post-period compared with the pre-period for all groups, i.e., office-based City employees, Department of Education employees, and non-City employees (Table 2).

**Table 2.**
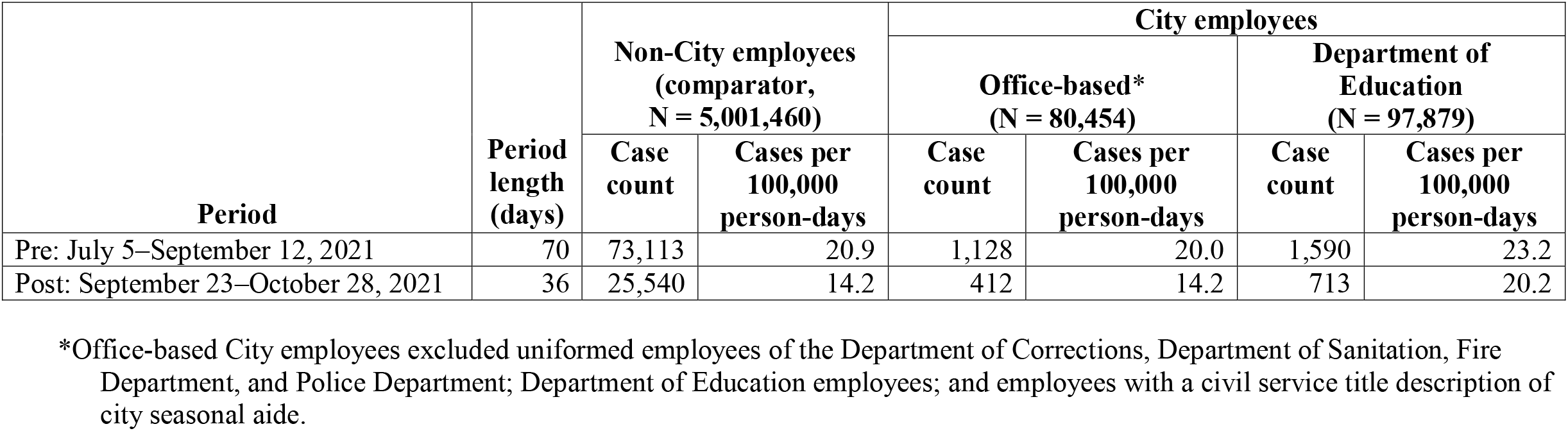
COVID-19 cases diagnosed among New York City residents 18–64 years-old before and after September 13, 2021, when the municipal employee return-to-office mandate was enacted and public schools reopened, by municipal employment status.

| Period | Period length (days) | Non-City employees (comparator, N = 5,001,460) |  | City employees |  |  |  |
| --- | --- | --- | --- | --- | --- | --- | --- |
|  |  |  |  | Office-based* (N = 80,454) |  | Department of Education (N = 97,879) |  |
|  |  | Case count | Cases per 100,000 person-days | Case count | Cases per 100,000 person-days | Case count | Cases per 100,000 person-days |
| Pre: July 5–September 12, 2021 | 70 | 73,113 | 20.9 | 1,128 | 20.0 | 1,590 | 23.2 |
| Post: September 23–October 28, 2021 | 36 | 25,540 | 14.2 | 412 | 14.2 | 713 | 20.2 |

\*Office-based City employees excluded uniformed employees of the Department of Corrections, Department of Sanitation, Fire Department, and Police Department; Department of Education employees; and employees with a civil service title description of city seasonal aide.

**Figure 1.**
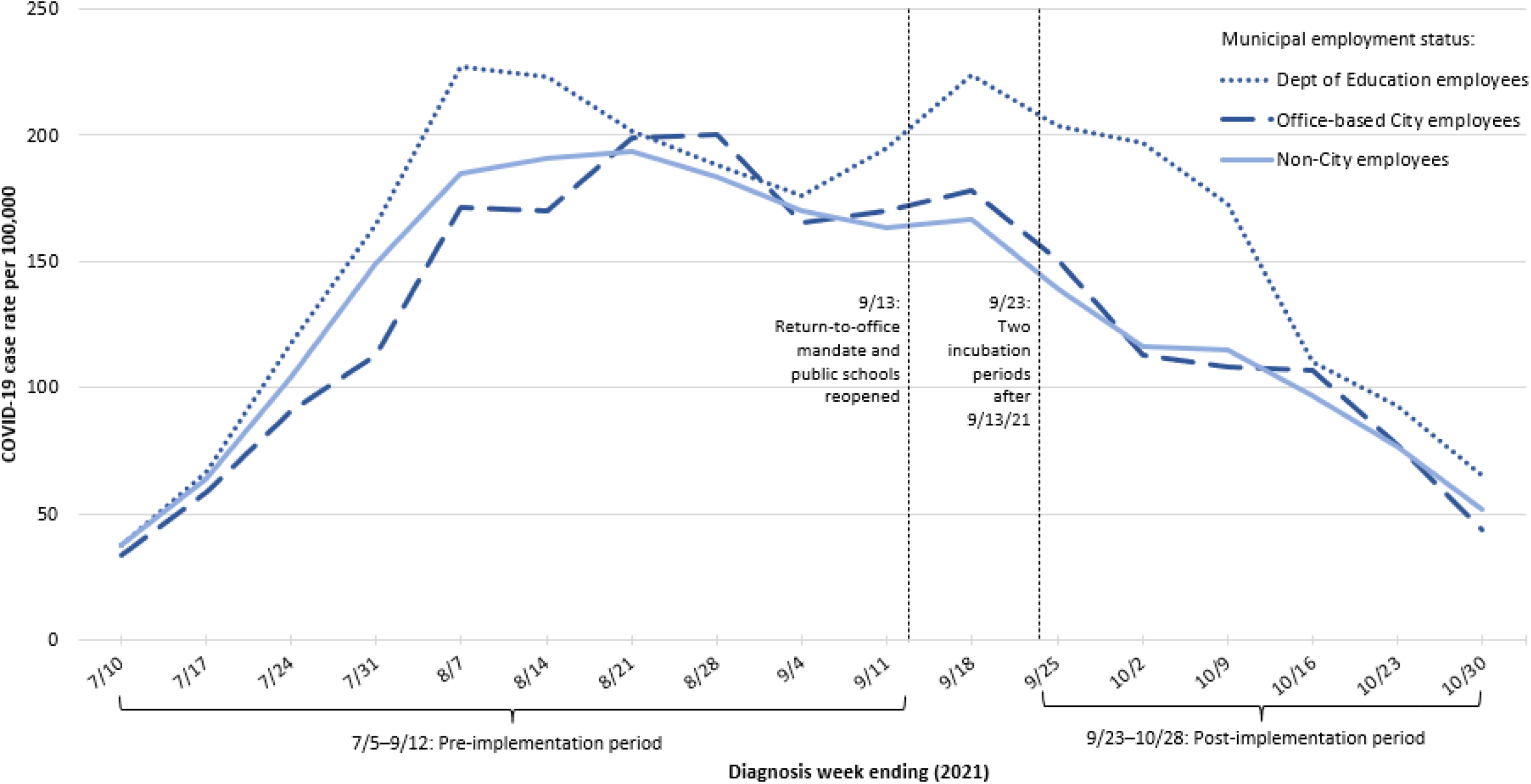
Weekly COVID-19 case rates among New York City residents 18–64 years-old, by municipal employment status, July 5– October 28, 2021* *The first and last weeks depicted are partial weeks: week ending July 10 reflects diagnoses July 5–10; week ending October 30 reflects diagnoses October 24–28.

#### Office-Based City Employees

Among non-City employees, we found a 32.1% (95% CI: 31.1%–33.0%) reduction in the COVID-19 case rate during the post-period compared with the pre-period. Among office-based City employees, we found a 29.0% (95% CI: 20.5%–36.6%) reduction in the case rate during the post-period compared with the pre-period. The incidence rate ratio among office-based City employees comparing the post-to pre-mandate periods was approximately the same (4.6% increase; 95% CI: 6.8% decrease–17.0% increase) as it would have been had the office-based City employees followed the same trend as non-City employees. Epidemic trends were similar between the two groups (Figure 1).

#### Department of Education Employees

Among Department of Education employees, we found only a 12.8% (95% CI: 4.8%– 20.2%) reduction in the case rate during the post-period compared with the pre-period. The incidence rate ratio among Department of Education employees comparing the post-to pre-mandate periods was 28.4% (95% CI: 17.3%–40.3%) larger than it would have been had Department of Education employees followed the same trend as non-City employees.

Among Department of Education employees, the proportion of COVID-19 cases diagnosed among employees with a civil service title of teacher increased from 44.7% (710/1,590) during the pre-implementation period to 57.5% (410/713) during the post-implementation period. The proportion of Department of Education case-patients who were known to be symptomatic with COVID-19–like illness also increased from the pre-implementation period (87.0%) to the post-implementation period (93.1%) (Table 1). During the washout period to account for worksite transmission to occur, case rates decreased among non-City employees and increased among Department of Education employees (Figure 1), possibly reflecting transmission stemming from staff returning to schools prior to students.

#### Non-Inferiority Tests

During the post-implementation period, the excess average weekly cases per 100,000 office-based City employees was 4.4 (95% CI: 3.0–5.6), which was within the *a priori* threshold consistent with “low” worksite transmission of <10. In contrast, the excess average weekly cases per 100,000 Department of Education employees was 31.3 (95% CI: 29.7–32.8), exceeding the threshold consistent with “low” worksite transmission (See Table, Supplemental Digital Content 4, which shows the logic for the non-inferiority tests).

### Vaccination Mandate

The Delta wave had plateaued in NYC when the vaccination mandate was implemented on October 29, 2021^37^ (Figure 2). Accordingly, the case rate per 100,000 person-days was similar during the post-implementation period for the primary analysis compared with the pre-implementation period among all groups, i.e., all City employees, employees of agencies with higher and lower vaccination rates, and non-City employees (Table 3A). Dramatic increases in COVID-19 case rates (Table 3A) and hospitalization rates (Tables 3B–3C) were observed during the post-implementation period for the secondary analysis, which extended through the large Omicron (BA.1) wave (see Figures, Supplemental Digital Content 5 and 6, which illustrate large increases in case rates and hospitalization rates, respectively, during January 2021 among all groups).

**Table 3.** COVID-19 cases and hospitalizations diagnosed among New York City residents 18–64 years-old before and after October 29, 2021, when the municipal employee vaccination mandate was enacted, by municipal employment status.

| Period | Period length (days) | Non-City employees (comparator, N=5,001,460) |  | City employees |  |  |  |  |  |
| --- | --- | --- | --- | --- | --- | --- | --- | --- | --- |
|  |  |  |  | All (N=212,953) |  | Employed at agency with higher vaccination coverage<sup>†</sup> (N=104,628) |  | Employed at agency with lower vaccination coverage<sup>‡</sup> (N= 66,574) |  |
|  |  | Case count | Cases per 100,000 person-days | Case count | Cases per 100,000 person-days | Case count | Cases per 100,000 person-days | Case count | Cases per 100,000 person-days |
| Pre: September 23–October 28, 2021 | 36 | 25,540 | 14.2 | 1,464 | 19.1 | 769 | 20.4 | 531 | 22.2 |
| Post (primary analysis):<br>October 29–November 30, 2021 | 33 | 28,019 | 17.0 | 1,284 | 18.3 | 690 | 20.0 | 431 | 19.6 |
| Post (secondary analysis):<sup>§</sup><br>October 29, 2021–January 31, 2022 | 95 | 753,196 | 158.5 | 39,516 | 195.3 | 19,476 | 195.9 | 16,284 | 257.5 |

B. COVID-19 hospitalizations (hosps), 18–64 year-olds
| Period | Period | Non-City employees (comparator, N=5,001,460) |  | City employees |  |  |  |  |  |
| --- | --- | --- | --- | --- | --- | --- | --- | --- | --- |
|  |  |  |  | All (N=212,953) |  | Employed at agency with higher vaccination coverage<sup>†</sup> (N=104,628) |  | Employed at agency with lower vaccination coverage<sup>‡</sup> (N= 66,574) |  |
|  |  | Hosp count | Hosps per 100,000 person-days | Hosp count | Hosps per 100,000 person-days | Hosp count | Hosps per 100,000 person-days | Hosp count | Hosps per 100,000 person-days |

| **Period** | **length**<br>**(days)** |  |  |  |  |  |  |  |  |
| --- | --- | --- | --- | --- | --- | --- | --- | --- | --- |
| Pre: September 23–October 28, 2021 | 36 | 970 | 0.54 | 35 | 0.46 | 9 | 0.24 | 20 | 0.83 |
| Post: October 29, 2021–January 31, 2022 | 95 | 14,985 | 3.15 | 309 | 1.53 | 167 | 1.68 | 98 | 1.55 |

C. COVID-19 hospitalizations (hosps), restricted to 50–64 year-olds
| **Period** | **Period**<br>**length**<br>**(days)** | **Non-City employees**<br>**(comparator,**<br>**N= 1,417,471)** |  | **City employees**<br>**(N=71,343)** |  |
| --- | --- | --- | --- | --- | --- |
|  |  | **Hosp**<br>**count** | **Hosps per**<br>**100,000**<br>**person-**<br>**days** | **Hosp**<br>**count** | **Hosps per**<br>**100,000**<br>**person-**<br>**days** |
| Pre: September 23–October 28, 2021 | 36 | 440 | 0.86 | 12 | 0.47 |
| Post: October 29, 2021–January 31, 2022 | 95 | 6,338 | 4.71 | 131 | 1.93 |

<sup>†</sup> Agencies with $\geq 96\%$ of employees with at least one COVID-19 vaccine dose as of October 30, 2021: Landmarks Preservation Commission, Office of Management and Budget, Mayor’s Office, Department of City Planning, Financial Information Services Agency/Office of Payroll Administration, Department of Consumer and Worker Protection, Department of Education, Department of Health and Mental Hygiene (including Office of Chief Medical Examiner), and Department of Small Business Services.<sup>47</sup>
<sup>‡</sup> Agencies with $< 90\%$ of employees with at least one COVID-19 vaccine dose as of October 30, 2021: Department of Corrections, Fire Department (including Emergency Medical Services), NYC Housing Authority, Department of Sanitation, NYC Employees Retirement System, Department of Homeless Services, Police Department, Department of Citywide Administrative Services, Department of Transportation, and Department of Probation.<sup>47</sup>
<sup>§</sup> Analyses to assess cases diagnosed during the first 3 months of the vaccination mandate (which included no requirement for a booster dose) were considered secondary because vaccine effectiveness against infection with the Omicron variant was reduced relative to prior variants, while vaccine effectiveness against severe illness remained high.

**Figure 2.**
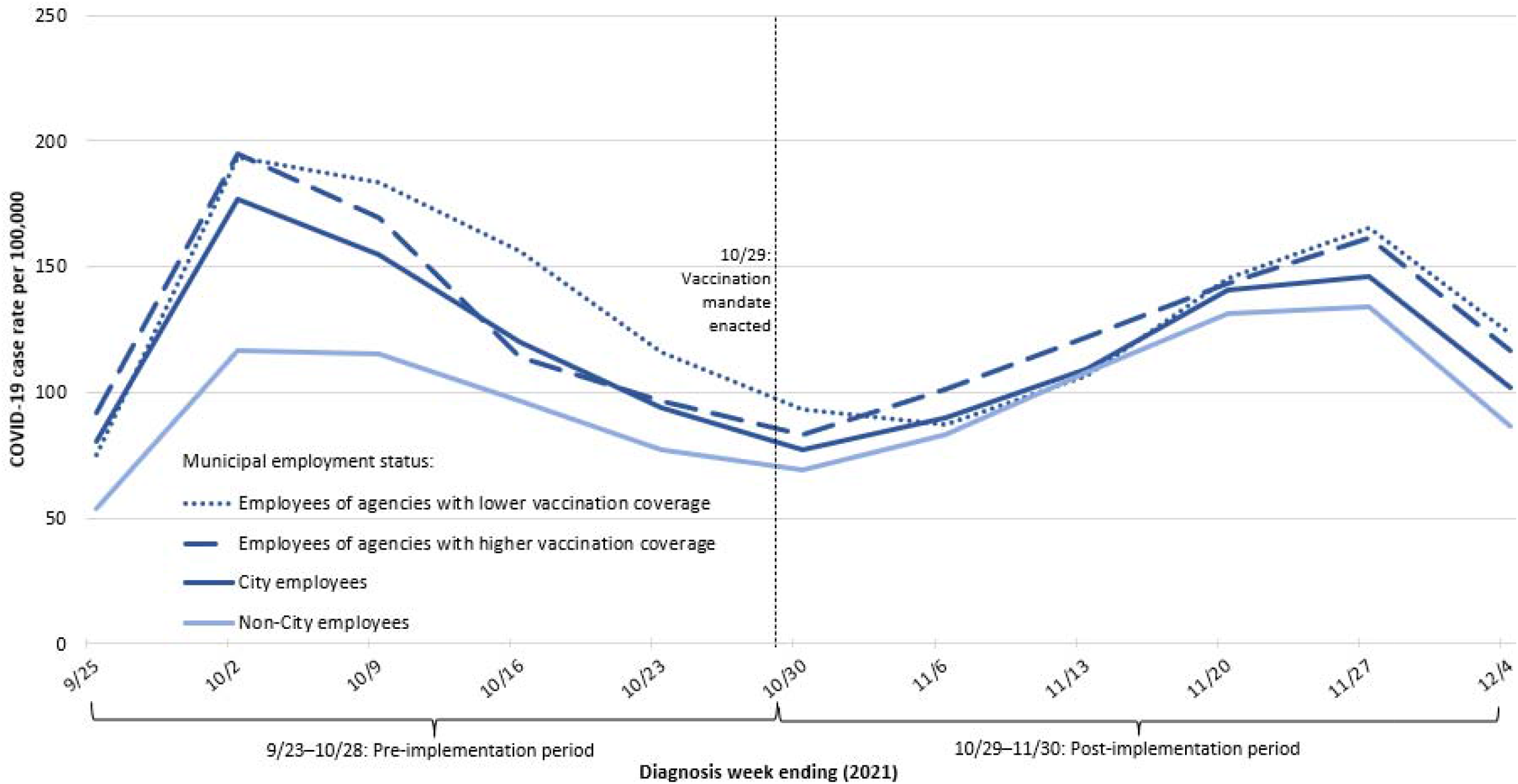
Weekly COVID-19 case rates among New York City residents 18–64 years-old, by municipal employment status, September 23–November 30, 2021.* *The first and last weeks depicted are partial weeks: week ending September 25 reflects diagnoses September 23–25; week ending December 4 reflects diagnoses November 28–30.

#### Primary Analysis

Among non-City employees, there was a 19.7% (95% CI: 17.7%–21.7%) increase in the COVID-19 case rate during October 29–November 30, 2021 (after implementation of the vaccination mandate and prior to Omicron emergence) compared with the pre-implementation period. Among City employees, there was little change in the case rate during the post-period compared with the pre-period (4.3% decrease; 95% CI: 11.2% decrease–3.1% increase). Among City employees, the incidence rate ratio comparing the post-period with the pre-period was 20.1% (95% CI: 13.7%–26.0%) smaller than it would have been had City employees followed the same trend as non-City employees.

#### Extended Post-Vaccination Mandate Implementation Period (October 29, 2021–January 31, 2022)

In a secondary analysis assessing the first 3 months of the vaccination mandate extending through the Omicron (BA.1) wave, non-City employees had a 11.2 (95% CI: 11.0–11.3)-fold increase in the case rate, while City employees had a 10.2 (95% CI: 9.7–10.8)-fold increase in the case rate during the post-period compared with the pre-period. Among City employees, the incidence rate ratio comparing the extended post-period with the pre-period was 8.5% (95% CI: 3.4%–13.2%) smaller than it would have been had City employees followed the same trend as non-City employees.

As for COVID-19 hospitalizations among the full study population of 18–64 year-olds, non-City employees had a 5.9 (95% CI: 5.5–6.3)-fold increase in the hospitalization rate, while City employees had a 3.3 (95% CI: 2.4–4.7)-fold increase in the hospitalization rate during the post-period compared with the pre-period. Among City employees, the hospitalization incidence rate ratio comparing the extended post-period with the pre-period was 42.9% (95% CI: 17.1%– 59.4%) smaller than it would have been had City employees followed the same trend as non-City employees.

We repeated this analysis, restricting to 50–64 year-olds as a subpopulation with greater underlying hospitalization risk. Older non-City employees had a 5.5 (95% CI: 5.0–6.0)-fold increase in the hospitalization rate, while older City employees had a 4.1 (95% CI: 2.3–7.5)-fold increase in the hospitalization rate during the post-period compared with the pre-period. Among older City employees, the hospitalization incidence rate ratio comparing the extended post-period with the pre-period was not statistically significantly different (24.2% smaller [95% CI: 56.6% smaller–45.5% larger]) than it would have been had older City employees followed the same trend as older non-City employees. This estimate should be interpreted with caution, as only 12 hospitalizations were observed among 50–64 year-old City employees during the 5-week pre-mandate period (Table 3C), leading to wide uncertainty. In addition, weekly hospitalization rates during the brief pre-mandate period appeared to be stable among older non-City employees but decreasing among older City employees (Supplemental Digital Content 6B), possibly violating the common trends assumption.

#### City Employees Stratified by Agency Vaccination Coverage

Another secondary analysis grouped employees of agencies with higher and with lower vaccination coverage and compared each group with non-City employees using a triple difference analysis. During the primary post-vaccination mandate period (October 29–November 30, 2021), City employees experienced decreased case rates compared with the pre-period: the decrease was 2.1% (95% CI: −8.5%–11.7%) among employees of agencies with higher vaccination coverage and 11.5% (95% CI: −0.5%–22.0%) among employees of agencies with lower vaccination coverage; as above, non-City employees experienced a 19.7% (95% CI: 17.7%–21.7%) increase. There was no statistically significant difference in case rates from the pre-to post-period when comparing employees of agencies with higher than lower vaccination coverage and controlling for the trend among non-City employees.

We repeated this analysis using the extended post-vaccination mandate implementation period that included the Omicron (BA.1) wave (October 29, 2021–January 31, 2022). The increase in case rates was 9.6 (95% CI: 8.9–10.3)-fold among employees of agencies with higher vaccination coverage and 11.6 (95% CI: 10.7–12.7)-fold among employees of agencies with lower vaccination coverage; as above, non-City employees experienced an 11.2 (95% CI: 11.0– 11.3)-fold increase. Controlling for the trend among non-City employees, the increase in case rates for employees of agencies with higher vaccination coverage was 17.4% (95% CI: 7.5%– 26.3%) lower than the increase in case rates for employees of agencies with lower vaccination coverage. The triple difference analysis was not conducted for the COVID-19 hospitalizations outcome because <10 hospitalizations were observed among employees of agencies with higher vaccination coverage during the pre-vaccination mandate implementation period (Table 3B), leading to unstable rates.

## DISCUSSION

The return-to-office mandate enacted on September 13, 2021 was not associated with a relative increase in COVID-19 cases diagnosed during September 23–October 28, 2021 among office-based municipal employees residing in NYC. The excess average weekly cases per 100,000 office-based City employees during the post-implementation period was low, at 4.4 (95% CI: 3.0–5.6). This finding supports that the safety measures in place during that period, including mandatory face coverings and proof of full vaccination or weekly negative PCR diagnostic tests, were effective in keeping worksite transmission low while the Delta variant predominated. This finding cannot be generalized to later periods because of the predominance of the more transmissible Omicron variant^48^ and the revised DCAS directive (March 7, 2022) authorizing City employees to remove their face coverings in the workplace.^49^

In contrast, the reopening of public schools for the 2021–2022 academic year was associated with an increase in COVID-19 cases among Department of Education employees, controlling for the trend among non-City employees. The excess average weekly cases per 100,000 Department of Education employees during September 23–October 28, 2021 was 31.3 (95% CI: 29.7–32.8), consistent with CDC’s definition at the time of “moderate” community transmission (i.e., 10.00–49.99 new cases per 100,000 persons in the past 7 days).^42^ This finding was consistent with contemporaneous local news reports of outbreaks affecting staff, related to congregating indoors with insufficient masking and social distancing.^50,51^ Additionally, unlike office-based City employees, Department of Education employees worked with children.

Asymptomatic SARS-CoV-2 infections are more common among children,^52^ so untested students could have unknowingly transmitted to staff. The finding of a relative short-term increase in COVID-19 cases among Department of Education employees cannot be generalized to later periods, as CDC expanded COVID-19 vaccination eligibility to 5–11 year-olds on November 2, 2021^53^ and to ≥6 month-olds–<5 year-olds on June 18, 2022,^54^ and school ventilation systems were further upgraded.^55,56^

COVID-19 vaccination mandates were previously demonstrated to increase vaccination coverage.^57,58^ Our analysis further demonstrated that the vaccination mandate for NYC municipal employees was associated with a relative decrease in COVID-19 cases. Case rates increased from the pre-to primary post-mandate implementation period through November 30, 2021 among non-City employees but not for City employees. In addition, despite reduced vaccine effectiveness against infection with the Omicron variant^45^ and no mandate for a booster dose, increasing trends among City employees from the pre-to secondary post-mandate implementation period through January 31, 2022 and extending through the Omicron (BA.1) wave were approximately 9% smaller for case rates and 43% smaller for hospitalization rates than they would have been had City employees followed the same trends as non-City employees. These findings support the effectiveness of the vaccination mandate in modestly reducing workplace transmission when Omicron (BA.1) predominated.

### Limitations

Difference-in-difference analyses rely on the common trends assumption that important unmeasured variables are either time-invariant group attributes or time-varying factors that are group-invariant.^36^ Specifically, we assumed that COVID-19 case ascertainment either (1) differed between City and non-City employees (due to, for example, differences in age distribution, prior SARS-CoV-2 infection history, or testing rates), but this difference was consistent over the study period, or (2) differed over the study period (due to, for example, epidemic trends^59^), but this difference was consistent for City and non-City employees. Figure 1 provides empirical support that during the 10 weeks prior to the first mandate, weekly trends in COVID-19 case rates for non-City employees, office-based City employees, and Department of Education employees were similar, as each group experienced an increase in cases during the Delta epidemic wave. City employees were also geographically representative of the general population (Supplemental Digital Content 2) and thus likely exposed to the same underlying epidemic trends. Other policies implemented during the study period likely affected City and non-City employees consistently, including on September 13, 2021 the start of enforcement of the Key to NYC vaccination mandate applying to patrons of gyms, restaurants, and indoor entertainment venues.^60^ Similarly, we would not expect that use of at-home rapid antigen tests, which became widely available in NYC starting mid-December 2021^61^ and were not reportable to DOHMH, would substantially differ by City employment status.

Differential testing requirements for City and non-City employees were the main threat to study validity. A weekly PCR diagnostic testing requirement for unvaccinated City employees^5^ (including unvaccinated Department of Education employees^9^) was in effect during the post-implementation period for the return-to-office mandate and reopening of public schools.

Similarly, the federal Occupational Safety and Health Administration issued an emergency temporary standard effective November 5, 2021–January 26, 2022 for weekly testing of employees who were not fully vaccinated, applying to employers with at least 100 employees.^62^ Mandatory routine testing for subsets of City and non-City employees at different times might have led to differential case ascertainment and biased our findings. Reason for testing (e.g., workplace requirement, pre-travel clearance, etc.) was incomplete for reported cases. However, the proportion of case-patients by municipal employment status who were known to be symptomatic with COVID-19–like illness did not vary markedly by timing of diagnosis (Table 1), suggesting the magnitude of this potential bias is likely small. Furthermore, among Department of Education employees, the proportion of case-patients who were known to be symptomatic with COVID-19–like illness increased following reopening of public schools, indicating that the relative increase in case rates cannot be explained by increased ascertainment of asymptomatic infections. We are unaware of other reasons that differences in COVID-19 case trends between City and non-City employees before and after the mandates could not be causally attributable^63^ to the mandates.

There were at least three additional limitations. First, as with any analysis of matched datasets, overmatching and undermatching are possible,^35^ resulting in misclassification of City employment status for some case-patients and likely biasing findings toward the null.

Undermatching was more likely for employees of agencies for whom home address was frequently unavailable due to security concerns, which included two of the agencies (Police Department, Department of Corrections) that also had lower vaccination coverage. However, two of the 12 primary matching keys and all five secondary matching keys used identifiers other than address information (Supplemental Digital Content 1). Second, the interventions were gradually and partially adopted. Prior to the full-time return-to-office mandate on September 13, 2021, some office-based City employees had already returned to the office on at least a part-time basis. Under the City of New York’s Equal Employment Opportunity Policy, eligible employees were entitled to reasonable accommodations to continue to telework full-time (e.g., due to a disability) and/or to be exempted from the vaccination requirement due to medical or religious reasons.^64^ Furthermore, as of October 28, 2021, the day before the vaccination mandate, 74.3% of NYC residents had already received at least one COVID-19 vaccine dose.^65^ Following the vaccination mandate, thousands of City employees who were non-compliant were placed on unpaid administrative leave,^66^ so would not have reported to office settings or contributed to worksite transmission.

Publicization of the municipal employee mandates might have influenced telework reductions and vaccination uptake for employees in other sectors. These phenomena would reduce differences between the intervention and comparator groups and thus likely bias findings toward the null.

Third, we assessed only the direct effects of municipal employee mandates on workplace transmission among NYC residents of working age. The mandates also might have indirectly affected transmission among the group at highest risk for severe COVID-19 illness, ≥65 year-olds, e.g., through employees residing in multigenerational housing.^67,68^ Indirect effects, such as increases or decreases in transmission from City employees to their household contacts relative to non-City employees to their household contacts, were not assessed.

## Conclusion

We found short-term evidence that the return-to-office and vaccination mandates for NYC municipal employees were successful with respect to minimizing workplace transmission among City employees and that reopening public schools was associated with a relative increase in COVID-19 cases among Department of Education employees. These findings should be interpreted in broader context with other health, social, educational, and economic effects of the mandates, including municipal workforce attrition.^20^

As of July 2022, 33% of employed U.S. adults (53% of education workers, 33% of white collar/office-based workers, and 24% of blue-collar workers) remained moderately or very concerned about being exposed to COVID-19 at work.^69^ The ability to assess longer-term effects of the NYC mandates on COVID-19 case trends is limited in part by wide adoption of at-home rapid antigen tests, which are not reportable to DOHMH.^70^ Guidance to prevent and reduce transmission in workplaces is available from public health authorities, including DOHMH^71^ and CDC.^72^

## Supporting information

Supplemental Digital Content

## Data Availability

Line-level data are not publicly available in accordance with patient confidentiality and privacy laws. Publicly available data are linked below.

https://www1.nyc.gov/site/doh/covid/covid-19-data.page

## Abbreviations

(CDC): U.S. Centers for Disease Control and Prevention
(CI): confidence interval
(DCAS): Department of Citywide Administrative Services
(DOHMH): Department of Health and Mental Hygiene
(NYC): New York City
(UHF): United Hospital Fund

## Acknowledgments

The authors thank the NYC municipal workforce for serving their communities through challenging pandemic circumstances. We thank Jessica Sell, MPH for contributions to study planning, Rebecca Kahn, PhD, MS for contributions to the analytic plan, and all DOHMH staff who served in the Surveillance and Epidemiology Section of the NYC Incident Command System.

The authors received no specific funding for this work beyond their usual salaries. Dr. Greene was supported by the Public Health Emergency Preparedness Cooperative Agreement (grant No. NU90TP922035-03-03), funded by the US Centers for Disease Control and Prevention.

