## Supplemental Digital Content for "Effects of return-to-office, public schools reopening, and vaccination mandates on COVID-19 cases among municipal employee residents of New York City"

**Supplemental Digital Content 1**. Text showing the steps for dataset cleaning, standardization, reformatting, and matching and the 17 keys used for the deterministic match of employee records and disease surveillance data.

**Supplemental Digital Content 2**. Figure that illustrates that United Hospital Fund neighborhoods with larger percentages of the general population tended to have larger percentages of New York City employees.

**Supplemental Digital Content 3**. Figure that illustrates that among New York City residents 18–64 years-old, City employees generally skewed older than non-City employees.

**Supplemental Digital Content 4**. Table showing logic for non-inferiority tests.

**Supplemental Digital Content 5**. Figure showing weekly COVID-19 case rates among New York City residents 18–64 years-old, by municipal employment status, September 23, 2021–January 31, 2022.

**Supplemental Digital Content 6**. Figure showing COVID-19 hospitalization rates by week of diagnosis among New York City residents, by municipal employment status, September 23, 2021–January 31, 2022. (A) 18–64 year-olds. (B) 50–64 year-olds.

**Supplemental Digital Content 1**

Dataset cleaning, standardization, reformatting, and matching

We converted all numeric variables to uppercase character variables; compressed and removed all spaces and blanks in character strings; created new variables based on the number of letters for the first, middle, and last name; created “soundex variables” for the first and last name to facilitate searching for phonetic similarity; created month of birth, day of birth, and year of birth from the birthdate variable; and created a composite string format variable for home address by combining address fields (e.g., street number, street name, and apartment number) and standardizing street name suffixes. Frequently reported worksite addresses for Police Department, Department of Corrections, and Department of Investigations employees were converted into missing values. Reformatting and standardization of matching variables and the matching process were conducted using SAS® version 9.4 (SAS Institute, Inc., Cary, North Carolina).

The matching process incorporated standardized and cleaned variables from input datasets by selecting only matching keys that had valid and non-missing variables. Records were linked using any matching key. This was a one-to-many match process because any City employee could have multiple COVID-19 diagnoses >90 days apart during the study period; thus, each DCAS record went through all keys, but a COVID-19 case after matching to a DCAS record was removed from the matching pool so that subsequent employees could not also match to that case.

Overall, the 12 primary match keys linked 45,126 records; primary keys 1 and 12 resulted in the highest and lowest number of matched records (n=40,293 and n=17, respectively). For records that could not be matched using the 12 primary keys, we conducted a secondary match using 5 secondary, less restricted keys. The secondary match keys linked 21,447 records for manual review using composite home address, through which 1,467 records were further identified as true matched records. In an evaluation after this process, we identified and corrected 9 falsely matched records.

Next, cases among 11,014 patients with worksite addresses for Police Department, Department of Corrections, and Department of Investigations employees were re-evaluated. Using the secondary match keys, 7,382 records were matched to the DCAS list and had a valid NYC residential address on any laboratory report and were included in analysis as City employees; the remaining 3,632 were excluded from analysis because NYC residency could not be assumed.

17 keys used for the deterministic match of employee records and disease surveillance data

*Primary keys*

1: full last name, first 6 letters of first name, date of birth (DOB), borough-block-lot (BBL), building identification number (BIN)

2: full last name, first 3 letters of first name, DOB, BBL, BIN

3: first 4 letters of last name, first 2 letters of first name, DOB, BBL, BIN

4: first letter of last name, letters 3 to 8 of last name, letters 2 to 8 of first name, DOB, BBL, BIN

5: letters 4 to 7 of last name, full first name, DOB, BBL, BIN

6: soundex* last name, letters 3 to 5 of first name, DOB, BBL, BIN

7: soundex* last name, soundex* first name, DOB, BBL, BIN

8: full last name, full first name, year of DOB, month of DOB, BBL, BIN

9: full last name, full first name, year of DOB, day of DOB, BBL, BIN

10: full last name, full first name, month of DOB, day of DOB, BBL, BIN

11: full last name, first 6 letters of first name, DOB, middle name

12: full last name, first 3 letters of first name, DOB, middle name

*Secondary keys for records that did not match in passes 1–12 (for manual review)*

13: full last name, first 6 letters of first name, DOB

14: full last name, first 3 letters of first name, DOB

15: first 4 letters of last name, first 2 letters of first name, DOB

16: first letter of last name, letters 3 to 8 of last name, letters 2 to 8 of first name, DOB

17: letters 4 to 7 of last name, full first name, DOB

*SOUNDEX function, SAS 9.4, SAS Institute, Cary, NC (<https://documentation.sas.com/doc/en/pgmsascdc/9.4_3.2/lefunctionsref/n1i9a3o4kciemhn1kpgutl20e4i0.htm>)

**Supplemental Digital Content 2**.

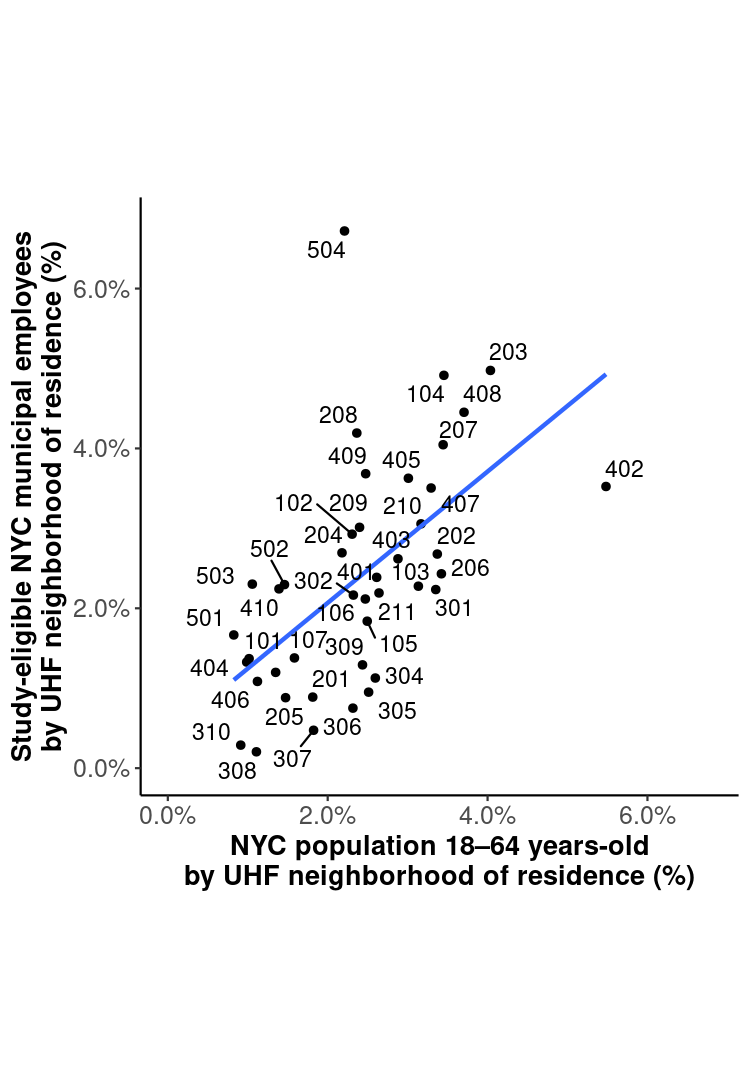

United Hospital Fund neighborhood map and ZIP code table available at: <https://www1.nyc.gov/assets/doh/downloads/pdf/ah/zipcodetable.pdf>.

**Supplemental Digital Content 3**

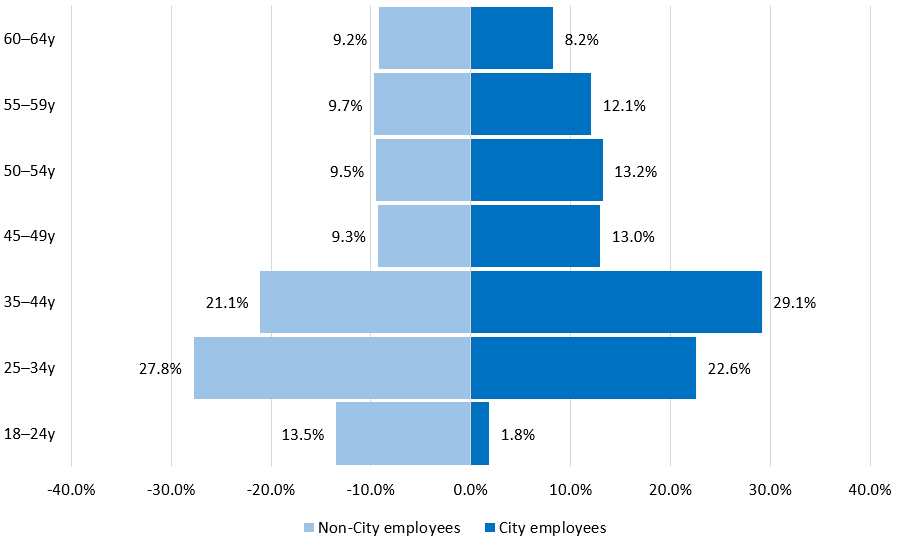

**Supplemental Digital Content 4**

Logic for non-inferiority tests to assess effects of return-to-office mandate and reopening of public schools.

|  | | | **Office-based City employees** | | **Department of Education employees** | |
| --- | --- | --- | --- | --- | --- | --- |
| **Step** | **Description** | **Source** | **Derivation** | **Result** | **Derivation** | **Result** |
| 1 | Observed^†^ average weekly COVID-19 cases per 100,000 employees during pre-implementation period | Table 2 | 1,128 cases / (80,454 office-based City employees * 70 observation days / 7 days per week / 100,000) | 140.2 | 1,590 cases / (97,879 Department of Education employees * 70 observation days / 7 days per week / 100,000) | 162.4 |
| 2 | Observed reduction in average weekly cases per 100,000 non-City employees from pre- to post-implementation periods | Difference-in-difference model | | 32.1%  (95% CI: 31.1%–33.0%) | Same as primary analysis | |
| 3 | Expected average weekly cases per 100,000 employees during post-implementation period, given observed reduction among non-City employees | Steps 1 and 2 | 140.2 *  (1 – 32.1%) | 95.2  (95% CI: 93.9–96.6) | 162.4 *  (1 – 32.1%) | 110.0  (95% CI: 108.8–111.9) |
| 4 | Observed^†^ average weekly cases per 100,000 employees during post-implementation period | Table 2 | 412 cases / (80,454 office-based City employees * 36 observation days / 7 days per week / 100,000) | 99.6 | 713 cases / (97,879 Department of Education employees * 36 observation days / 7 days per week / 100,000) | 141.6 |
| 5 | Excess average weekly cases per 100,000 employees during post-implementation period | Steps 3 and 4 | 99.6 – 95.2 | 4.4  (95% CI: 3.0–5.6) | 141.6 – 110.0 | 31.3  (95% CI: 29.7–32.8) |
| 6 | A priori threshold for excess average weekly cases per 100,000 employees during post-implementation period | Christie et al. 2021, [Table](https://www.cdc.gov/mmwr/volumes/70/wr/mm7030e2.htm?s_cid=mm7030e2_w#T1_down)  (DOI: [10.15585/mmwr.mm7030e2](http://dx.doi.org/10.15585/mmwr.mm7030e2)) | Low community transmission level | <10 | Same as primary analysis | |
| 7 | Test hypothesis that excess average weekly cases per 100,000 employees during post-implementation period was not ≥10: if lower bound of 95% CI for excess rate <10, then fail to reject null hypothesis; else if ≥10, then reject null hypothesis. | Steps 5 and 6 | 3.0 < 10 | Fail to reject null hypothesis that return-to-office mandate was not associated with a relative increase in COVID-19 cases among office-based City employees | 29.7 > 10 | Reject null hypothesis that reopening public schools was not associated with a relative increase in COVID-19 cases among Department of Education employees |

†Observed case counts were assumed to be known without error. Reference: Thygesen LC, Ersboll AK. When the entire population is the sample: strengths and limitations in register-based epidemiology. Eur J Epidemiol. 2014;29(8):551-8.

**Supplemental Digital Content 5**.

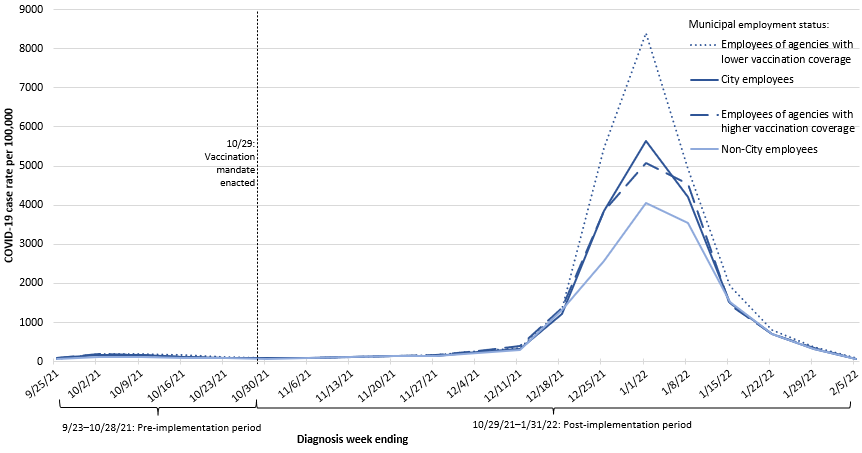

**Supplemental Digital Content 6A**.

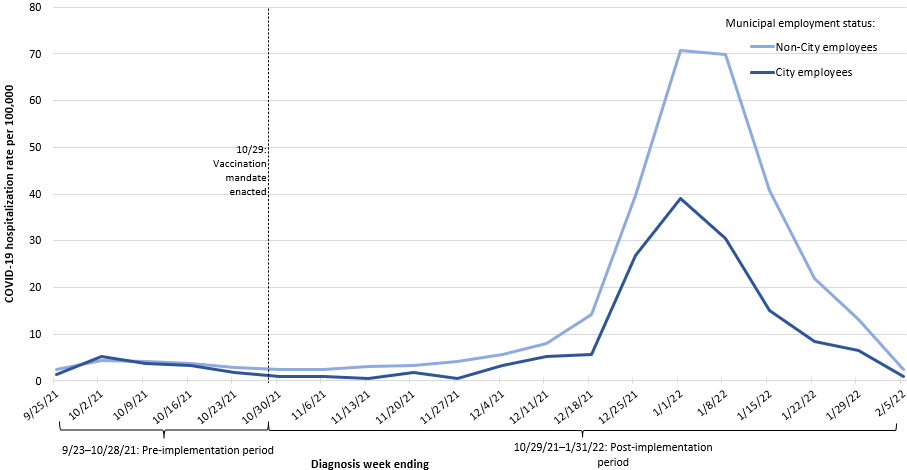

**Supplemental Digital Content 6B**.

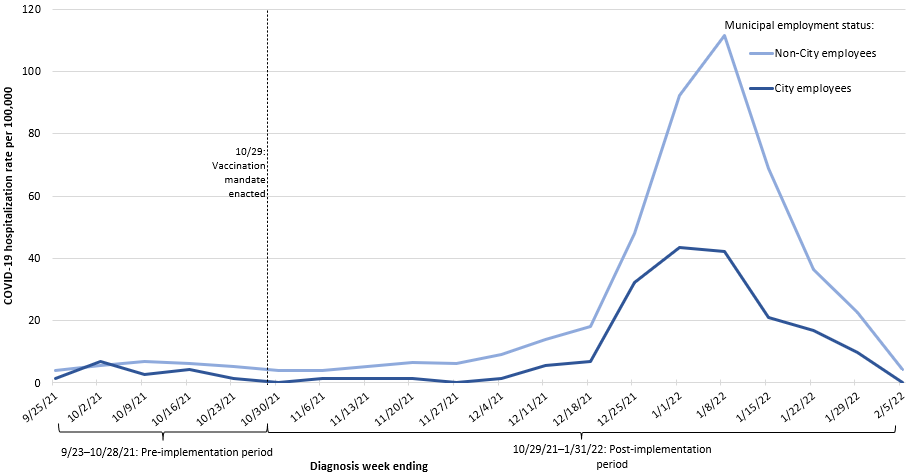
